# Lack of evidence for infectious SARS-CoV-2 in feces and sewage

**DOI:** 10.1101/2021.05.11.21256886

**Authors:** Sandra Albert, Alba Ruíz, Javier Pemán, Miguel Salavert, Pilar Domingo-Calap

**Author notes:** corresponding author: Pilar Domingo-Calap.

## Abstract

**Purpose:** The SARS-CoV-2 coronavirus is a respiratory virus whose primary route of transmission is airborne. However, it has been shown that the virus can replicate in gastrointestinal cells, can be excreted in feces, and can reach sewage systems. Although viral RNA has been found in patient feces and sewage, little is known about the potential fecal-oral transmission of the coronavirus. Determining the presence of infective viral particles in feces and sewage is necessary to take adequate control measures and to discover new routes of coronavirus transmission.

**Methods:** Feces and urine of COVID-19 patients, and wastewater samples at the time of high prevalence in the region under study (Valencia, Spain), have been analyzed both by molecular methods and cell culture.

**Results:** Presence of SARS-CoV-2 in feces of COVID-19 patients has been detected, even in patients without gastrointestinal symptoms, suggesting that viral shedding though stool is common. In addition, we have developed a sample concentration methodology that allows us to maintain the infectivity of the viral particles present in the samples. Finally, inoculation of cell cultures with fecal and sewage concentrated samples do not evidence the presence of infective viral particles.

**Conclusion:** There is no evidence of the presence of infectious SARS-CoV-2 in feces and sewage, suggesting that fecal-oral transmission is not a primary route. However, larger-scale efforts are needed to elucidate whether the fecal-oral transmission should be considered, especially with the emergence of new viral variants.

## Introduction

Severe acute respiratory syndrome coronavirus 2 (SARS-CoV-2) has caused a pandemic affecting the entire world. Although the main transmission route is via droplets or aerosols [1], it has been shown that the virus can replicate in intestinal mucosa [2], suggesting that viral excretion via feces could result in fecal-oral transmission of SARS-CoV-2 [3, 4]. Despite the efforts to isolate viral infectious particles from feces, little is known about the possibility of infecting new hosts via feces. In a first approach, it is necessary to determine the proportion of patients excreting virus in stool by analyzing fecal samples and testing them by molecular methods to detect viral RNA. In a second step, positive samples should be used to inoculate target cells expressing angiotensin-converting enzyme 2 (ACE2), the main receptor for SARS-CoV-2 [5], to determine the presence of infectious viruses in stool. Viral shedding in stool has been suggested to be more prolonged than in the respiratory tract, up to 5 weeks after respiratory samples have tested negative for SARS-CoV-2 RNA, even in mild or non-symptomatic patients [6, 7], including children [8].

The presence of SARS-CoV-2 RNA in feces has interesting applications, such as the implementation of new detection techniques through anal swabs (Sun et al. 2020), or epidemiological surveillance of SARS-CoV-2 through wastewater [10]. However, further efforts should be made to determine the proportion of patients shedding virus through feces, viral load per individual, and other parameters in order to use this specimen as a monitoring tool to correlate with incidence in the population. Therefore, it is important to determine whether positive specimens contain infectious particles, as only a few case reports have shown that viral infectious particles are found in stool [11–13]. Recently, fecal-oral transmission has been proposed as the most plausible explanation for two independent outbreaks in China, suggesting viral spread vis sewage aerosols in a building [14], and via sewage in a residential home [15]. In both cases, SARS-CoV-2 RNA sequencing supported the hypotheses. However, further efforts should be done to assess the fecal-oral route in SARS-CoV-2, and provide data based on the presence of infectious viral particles confirming the new route of transmission [12].

Here we propose the first attempt to evaluate the fecal excretion of SARS-CoV-2 in Spain, and the presence of viral infectious particles in feces, in a single-center study based on a small cohort of COVID-19 patients. In addition, to determine the possibility of the fecal-oral route, SARS-CoV-2 positive wastewater samples from Wastewater Treatment Plants of Valencia (Spain) were used to study the presence of viral particles.

## Materials and Methods

### Human subjects

This study has been approved by the ethics committee of the Instituto de Investigación Sanitaria La Fe (Valencia, Spain) to use patient samples, including urine and feces. In total, eight mild-symptomatic patients admitted to the Hospital Universitariy Politècnic La Fe between June and December 2020 we enrolled in this study. Informed consent was obtained from each patient. The criterion for inclusion in the study was to be over 18 years of age. Diagnosis and data collection from medical records were used. RT-qPCR on nasopharyngeal swabs was performed at the hospital following internal guidelines using different commercial available kits (BioFire® Respiratory Panel 2.1 (BioFire Defense LLC and BioFire Diagnostics LLC), Alinity m SARS-CoV-2 assay (Abbott Molecular), and Simplexa™ COVID-19 Direct (DiaSorin Molecular LLC)). Unfortunately, we were not able to recover the Ct values from nasopharyngeal swabs. Stool and urine samples were both suspended in Sigma Virocult viral transport medium without viral inactivators (W951S, MWE medical wire) and kept at 4°C until laboratory handling within 24 hours.

### Sewage samples

Wastewater from the metropolitan Wastewater Treatment Plants of Valencia (Spain) belonging to the Empresa Pública de Saneamiento de Aguas Residuales (EPSAR, Generalitat Valenciana) was sampled from December 2020 to January 2021. Grab and four-hour composite samples of 200 mL of sewage were collected in the morning. Samples were kept at 4 °C until they were processed in the laboratory within 24h of collection.

### Sewage sample concentration

Sewage samples were treated to allow viral particle concentration. Three different methods were tested: flocculation, ultra-filtration, and high-speed centrifugation. As a first common step, 10^5^ particle forming units (PFU) of the transmissible gastroenteritis virus (TGEV) were added to the wastewater samples as a surrogate virus. This was followed by gentle centrifugation at 3,000 × g, 10 min, at 4°C, and the supernatant was filtered through a 0.45 µm pore. Aluminum-driven flocculation was done in 50 mL of sewage, adjusting to pH 6.0, and adding 1:100 v:v of 0.9 N AlCl3 solution. Al(OH)3 precipitate was formed and after pH readjustment to 6.0, samples were gently agitated for 15 min at room temperature, and spun at 1,700 × g for 20 min. Pellets were resuspended into 10 mL of 3% beef extract (pH 7.4), and shaken for 10 min at 150 rpm. Samples were then centrifuged at 1,900 × g for 30 min and the pellet was resuspended in 1 mL of phosphate buffer (PBS) (Randazzo et al., 2020). Ultra-filtration method was done in 20 mL of sewage. Amicon tubes with 100KDa filters were used, and after 3,200 × g, 30 min, at 4°C, the residual liquid in the upper part of the filter was collected. High-speed centrifugation method was done in 35 mL of filtered wastewater, centrifuged at 80,000 × g, 3 h 30 min, at 4°C. The supernatant was removed and the pellet was resuspended in 500 µL of PBS.

### Processing of fecal samples

Fecal samples were resuspended in PBS to a final volume of 2 mL. Samples were then centrifuged at 3220 × g, 10 min, at 4°C twice to recover viruses in the supernatant. The supernatants were filtered through a 0.45 µm pore.

### RNA extraction and RT-qPCR

RNA extraction was performed with the Nucleospin RNA Virus Kit (Macherey-Nagel) following the recommended protocols. RT-qPCR was performed with the GoTaq® Probe 1-Step RT-qPCR System (Promega) using the U.S. Center for Disease Control N1 and N2 primers sets (2019-nCoV CDC EUA Kit, 1000rxn) provided by IDT (Integrated DNA Technologies). For each RT-qPCR run, calibration curves were performed using the 2019-nCoV_N_Positive Control provided by IDT. In addition, positive and negative controls (concentration, extraction and PCR) were included. To estimate viral loads, cycle threshold (Ct) values were used to calculate genomic copies (gc) per liter in the original sample due to calibration curves. Ct values bellow 40 were considered positive for SARS-CoV-2, as previously proposed [16]. All the experiments were performed in duplicate. In addition, and as an internal control, in-house TGEV primers were used to evaluate the efficacy of the protocol.

### High-throughput sequencing

Genomic sequencing was performed following the protocol of retrotranscription and amplification of ARTIC, using the amplification protocol, using the V3 version scheme for multiplex PCRs. This was followed by the preparation of libraries using the Nexus kit using the Nextera Flex kit (Illumina) and their sequencing on the Illumina MiSeq platform (paired-end reads 2×200 bp) [17].

### Cell culture

Fecal specimens and sewage samples with highest RNA detected by RT-qPCR were used to inoculate Vero E6 cells. To maintain infectivity, the high-speed centrifugation method was used to concentrate viruses in positive samples, as previously tested with TGEV.

Sewage samples were centrifuged at 3000 × g, 10 min, at 4°C and the supernatant was filtered through a 0.45 µm pore. Thirty-five mL of filtrate was centrifuged at 80,000 × g, 3h 30min, at 4°C and the pellet was resuspended in 500 µL of Dulbecco Modified Eagle Medium (DMEM) supplemented with 2% fetal bovine serum, non-essential amino acids, penicillin and streptomycin.

Fecal samples were centrifuged 3h 30min at 20,000 × g at 4°C to recover viruses, which were resuspended in 400 µL of Dulbecco Modified Eagle Medium (DMEM) supplemented with 2% fetal bovine serum, non-essential amino acids, penicillin and streptomycin, and 25 mM Hepes. All samples were analyzed undiluted and at 1/10 dilution. Diluted samples were used to avoid potential sample inhibitors or cytotoxic molecules. Negative controls were added in all experiments. For sewage, a negative RT-qPCR sample sampled in July 2020 was used as a negative control. For feces, a canine fecal sample was used as a negative control. Vero E6 cells were inoculated at 70% confluence with diluted and undiluted fecal or wastewater samples. Cultures were incubated at 37°C and 5% CO2 for six dpi. Replication of SARS-CoV-2 in cells cultures was analyzed by RT-qPCR, at 0 hpi, 48 hpi and 6 dpi. 100 µL of the supernatant was recovered and treated with DNA/RNA Shield (Zymo Researh) for each time point to inactivate the viruses and proceed to RNA extraction and RT-qPCR as explained above. In addition, microscopy images were taken to detect cytopathic effects of cell monolayers. All the experiments were performed in triplicate.

## Results

### Patients

This study was performed on a cohort of eight COVID-19 patients admitted to the Hospital Universitari i Politècnic La Fe (Valencia, Spain) between June and December 2020. In our cohort, 5 patients of the 8 showed respiratory symptoms, and only 3 had gastrointestinal (GI) symptoms. Other common symptoms, such as fever and headache, were very frequent in patients. None of the patients included in our cohort were admitted to intensive care units. Fecal and urine samples were collected from the patients to determine the presence of SARS-CoV-2, and in some of them we had access to longitudinal samples (Table 1).

**Table 1.** Main characteristics of the COVID-19 cohort included in the study. PCR + nasopharyngeal swab to validate SARS-CoV-2 infection. GI: gastrointestinal.

| Patient code | Gender | Admission date | Fever | Headache | Odinophagy | Respiratory symptoms | Anosmia | Dysgeusia | GI symptoms |
| --- | --- | --- | --- | --- | --- | --- | --- | --- | --- |
| 1 | Male | Jun 2020 | NO | YES | YES | NO | NO | NO | NO |
| 2 | Male | Jun 2020 | YES | YES | NO | NO | NO | NO | NO |
| 3 | Male | Jun 2020 | YES | YES | NO | NO | NO | NO | NO |
| 4 | Male | Jun 2020 | YES | YES | NO | YES | NO | NO | NO |
| 5 | Female | Jul 2020 | YES | YES | YES | YES | NO | NO | YES |
| 6 | Male | Sep 2020 | YES | YES | NO | YES | NO | NO | YES |
| 7 | Male | Nov 2020 | YES | YES | NO | YES | NO | NO | NO |
| 8 | Male | Dec 2020 | NO | YES | YES | YES | NO | NO | YES |

### Sample concentration comparison

Different sample concentration methods were tested to select a protocol capable of concentrating the virus while maintaining viral infectivity, using the transmissible gastroenteritis coronavirus (TGEV) as model. The three methods tested were: ultrafiltration, aluminum-driven flocculation, and high-speed centrifugation. The concentration factor was obtained by RTqPCR using in-house primers and probes, and by plaque assay infecting swine testicular (ST) cells. Six replicates were performed per method. For maintaining infectivity, high-speed centrifugation was the best method (concentration factor: 31.29 ± 6.49), followed by ultrafiltration (concentration factor: 7.38 ± 1.26). In contrast, infectivity was impaired by aluminum-driven flocculation (concentration factor: 0.02 ± 0.02). As for RNA detection, high-speed centrifugation obtained similar results to the aluminum-driven flocculation method (concentration factor: 14.98 ± 0.91 vs. 16.42 ± 2.13), whereas ultrafiltration was not suitable (concentration factor: 2.85 ± 0.87). For these reasons, the high-speed centrifugation method was chosen as the best in terms of concentration and maintenance of viral infectivity.

### Detection of SARS-CoV-2 RNA in fecal and urine samples

The presence of SARS-CoV-2 in the patient samples was evaluated by RT-qPCR in urine and feces. Internal controls using the surrogate TGEV coronavirus were used in all the samples, validating the efficacy of our protocol (data not shown). The results showed that 6 of the 8 patients had viral excretion in feces in at least one of the samples analyzed, ranging from 8.34 × 10^3^ gc/L to 6.74 × 10^6^ gc/L (Table 2). One of the negative patients, it is worth mentioning that he was negative despite suffering from GI symptoms. The absence of viral shedding in this patient was not expected, and a possible explanation is that the sample was taken one month after the onset of the first symptoms, suggesting that perhaps earlier sampling might have different results. In parallel, urine samples were collected for patients to test for RNA detection. In our dataset, one patient showed viral shedding in urine in two consecutive samples, despite the low viral load found (ranging from 5.60 × 10^4^ gc/L to 1.78 × 10^5^ gc/L).

**Table 2.**
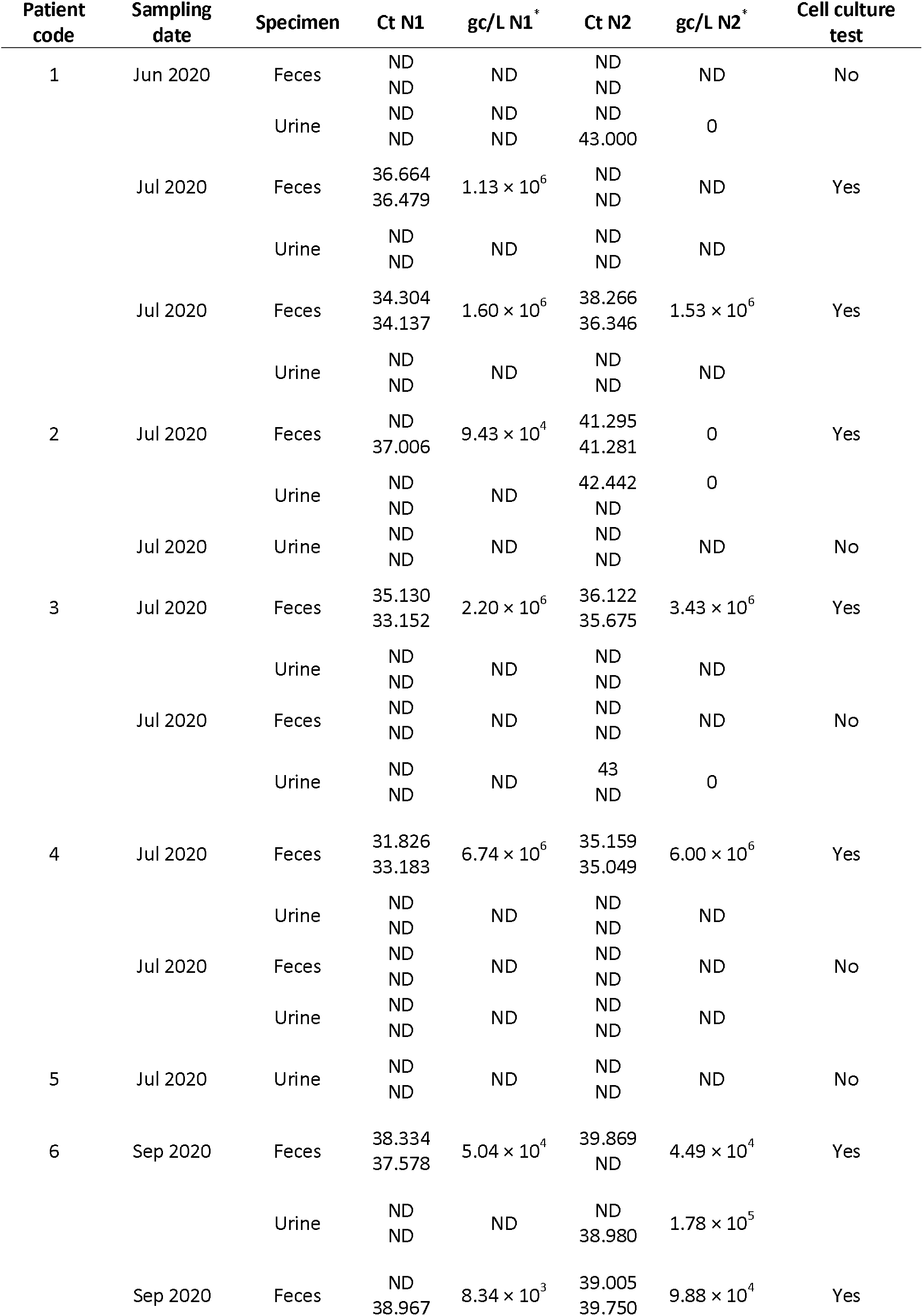
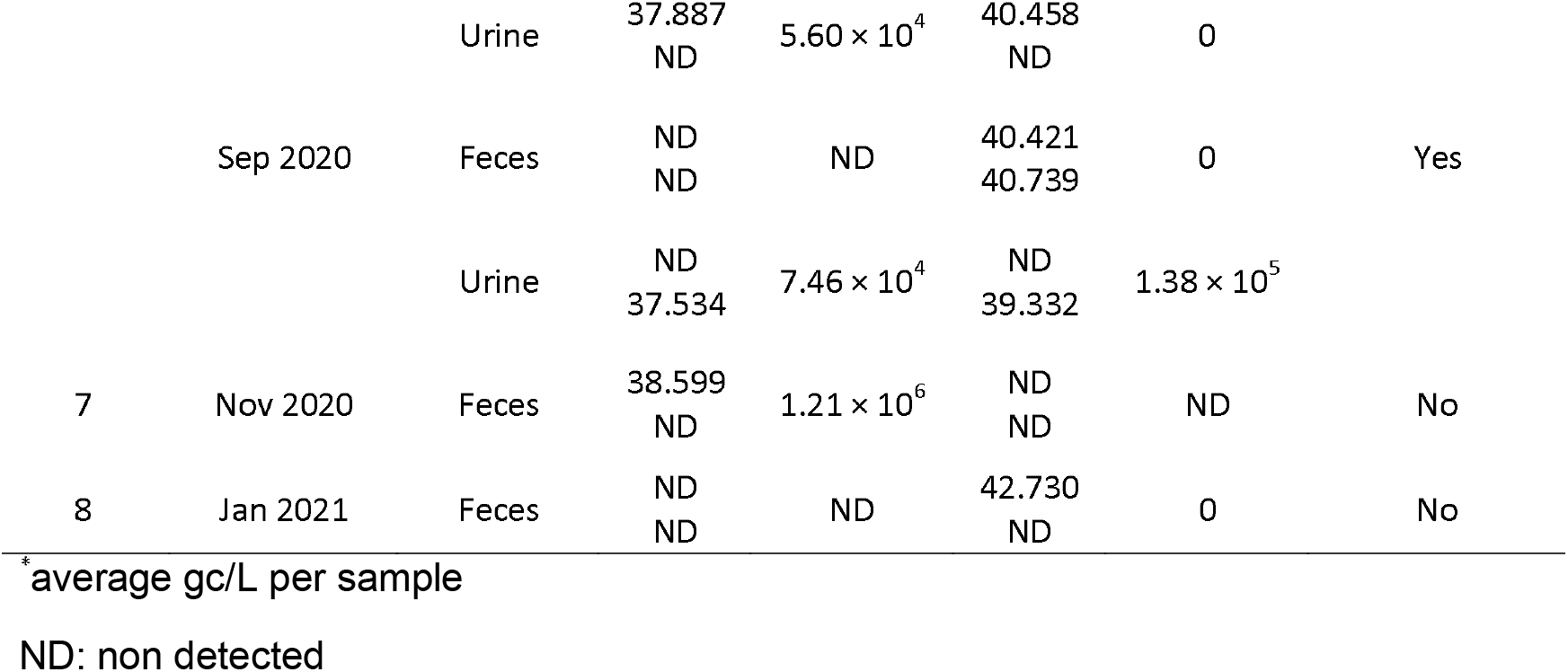
Detection of SARS-CoV-2 RNA in feces and urine of COVID-19 patients by RT-qPCR.

### SARS-CoV-2 sequencing attempt in fecal samples

In addition, we wanted to sequence the RNA obtained in the feces to analyze the variability found in stool. Unfortunately, the viral loads obtained in the feces of the patients tested were too low for viral sequencing. Despite the low copy number per mL in our samples, we attempted to sequence the sample with highest viral load, but the results were insufficient for further analysis. Using the Illumina MiSeq sequencer, and following the ARTICV3 + Nextera Flex protocol, only 13.28% of the SARS-CoV-2 genome was covered, with a coverage of 2 (unpublished data). Our results are consistent with previous data in which samples with low viral load are difficult to sequence [18]. Since the Ct values of the other samples were higher than those of the tested sample, we decided not to continue with the sequencing protocol for the remaining samples.

### Potential contribution of fecal shedding to SARS-CoV-2 transmission

To determine whether RNA found in feces was capable of infecting new hosts, we attempted to determine by cell culture the presence of infectious SARS-CoV-2 particles in positive samples, using 8 samples from 5 patients. Vero E6 cells were used as recommended cell models for SARS-CoV-2 infection [19]. Isolation of viral particles from feces and inoculation of cell monolayers were performed. Three sampling points were analyzed, 0 hours post-infection (hpi), 48 hpi and 6 days post-infection (dpi). No cytopathic effect on Vero E6 cells was observed in any of the samples analyzed (Figure 1). However, supernatants were collected and RT-qPCR was performed to detect viral amplification. In all samples tested no SARS-CoV-2 RNA was detected, suggesting the absence of viral particles in the cultures tested, despite the RNA found in the fecal samples (Table 3).

**Figure 1.**
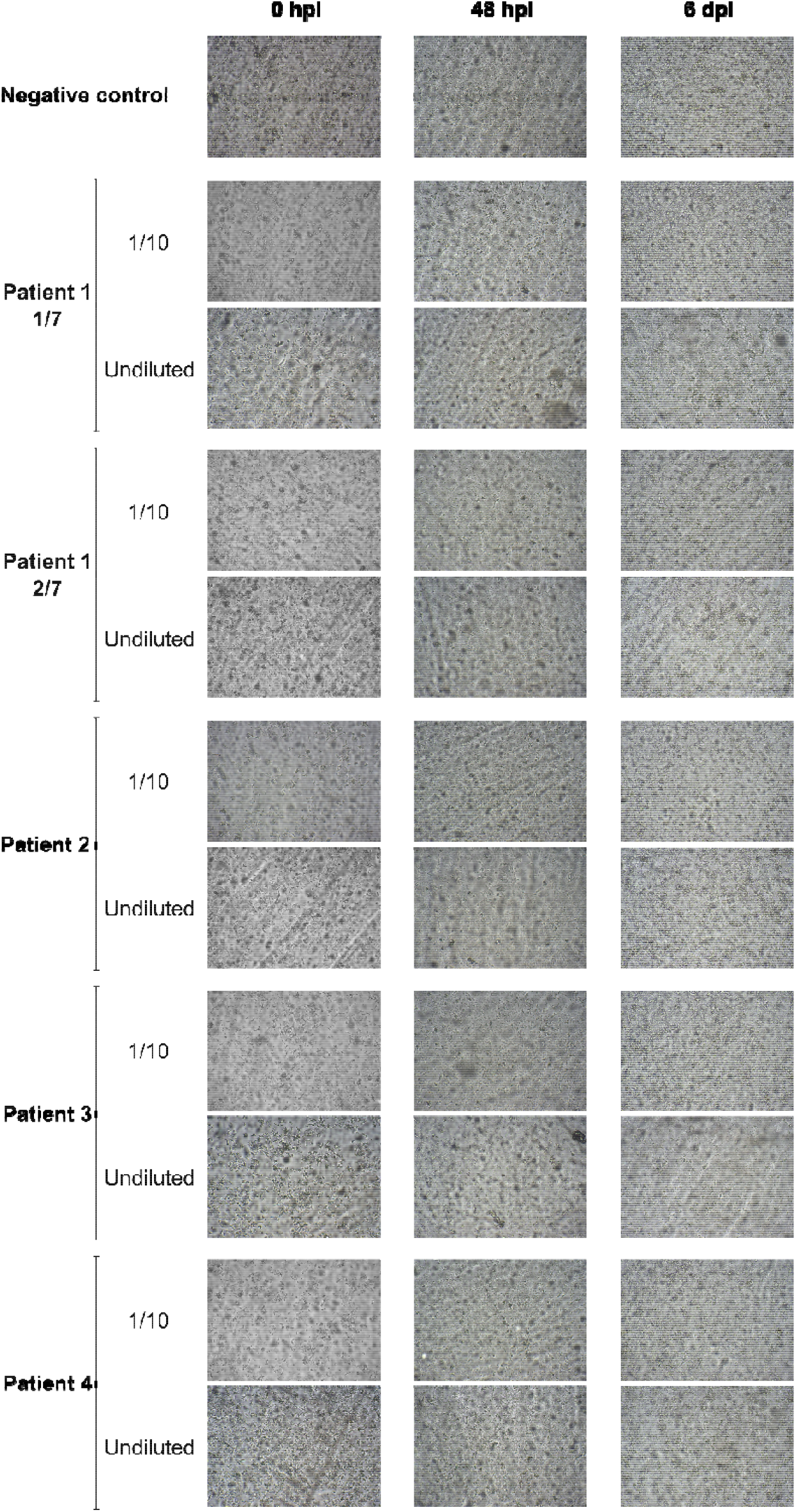
Representative images of Vero E6 cultures obtained at different time points inoculated with fecal-treated samples. Undiluted and 1/10 diluted samples were tested for each sample. Magnification: 40X. In patient 1, 1/7 and 2/7 refer to two different samples.

**Table 3.**
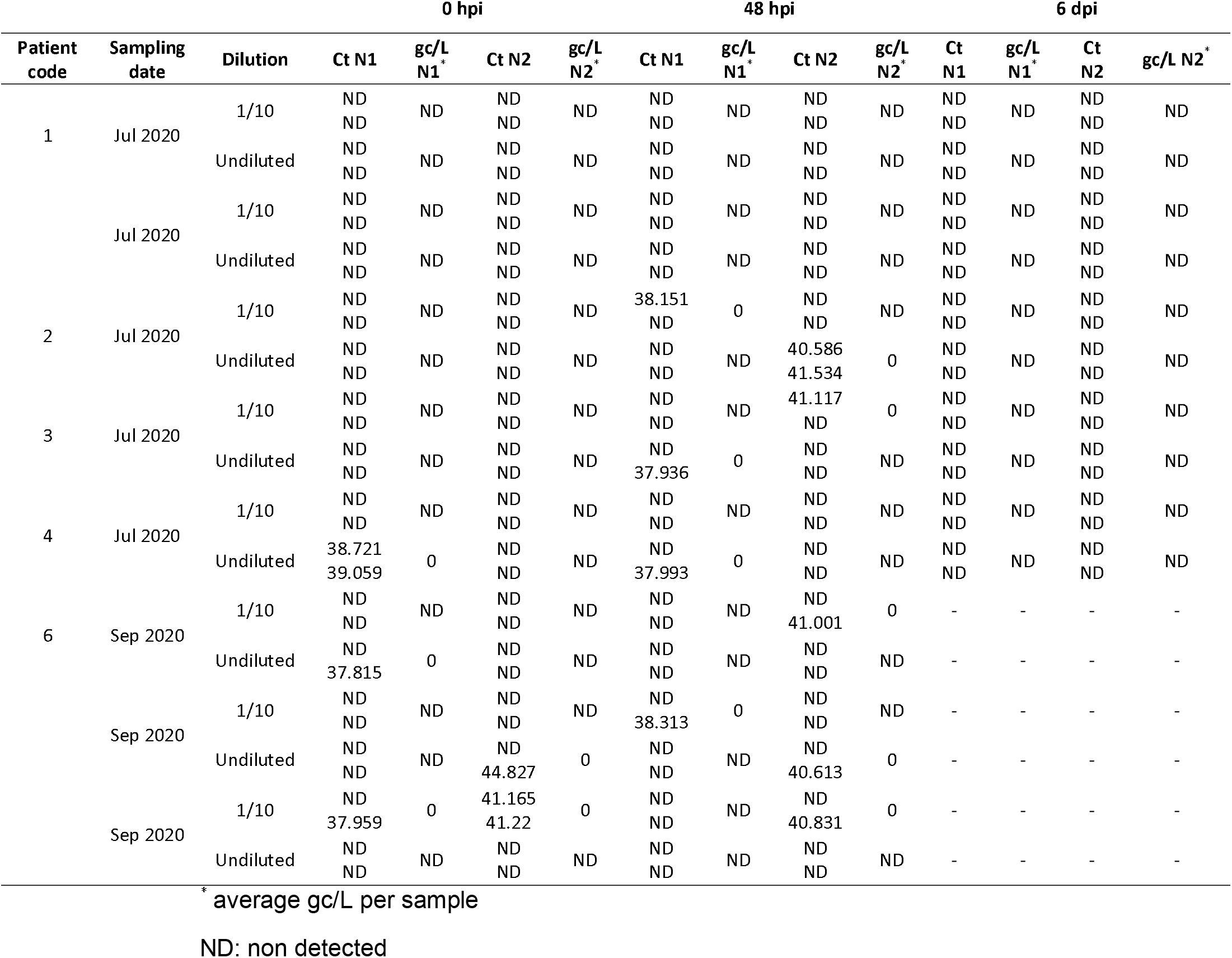
Detection of SARS-CoV-2 RNA in inoculated Vero E6 cultures with treated feces of COVID-19 patients by RT-qPCR after 0 hpi, 48 hpi and 6 dpi.

### Potential contribution of wastewater to SARS-CoV-2 transmission

Further, we wanted to determine the potential contribution of wastewater to SARS-CoV-2 transmission. Eight sewage samples from wastewater treatment plants in Valencia (Spain) between December 2020 and January 2021, where the incidence rate in the target population was higher than 1000 per 100000 people. Samples were concentrated by high-speed centrifugation to ensure virus viability and analyzed by RT-qPCR and cell culture. Again, despite the high viral load obtained in the samples (ranging from 1.43 × 10^4^ gc/L to 1.48 × 10^6^ gc/L) (Table 4), we were unable to detect evidence of replication in our assays (Table 5).

**Table 4.**
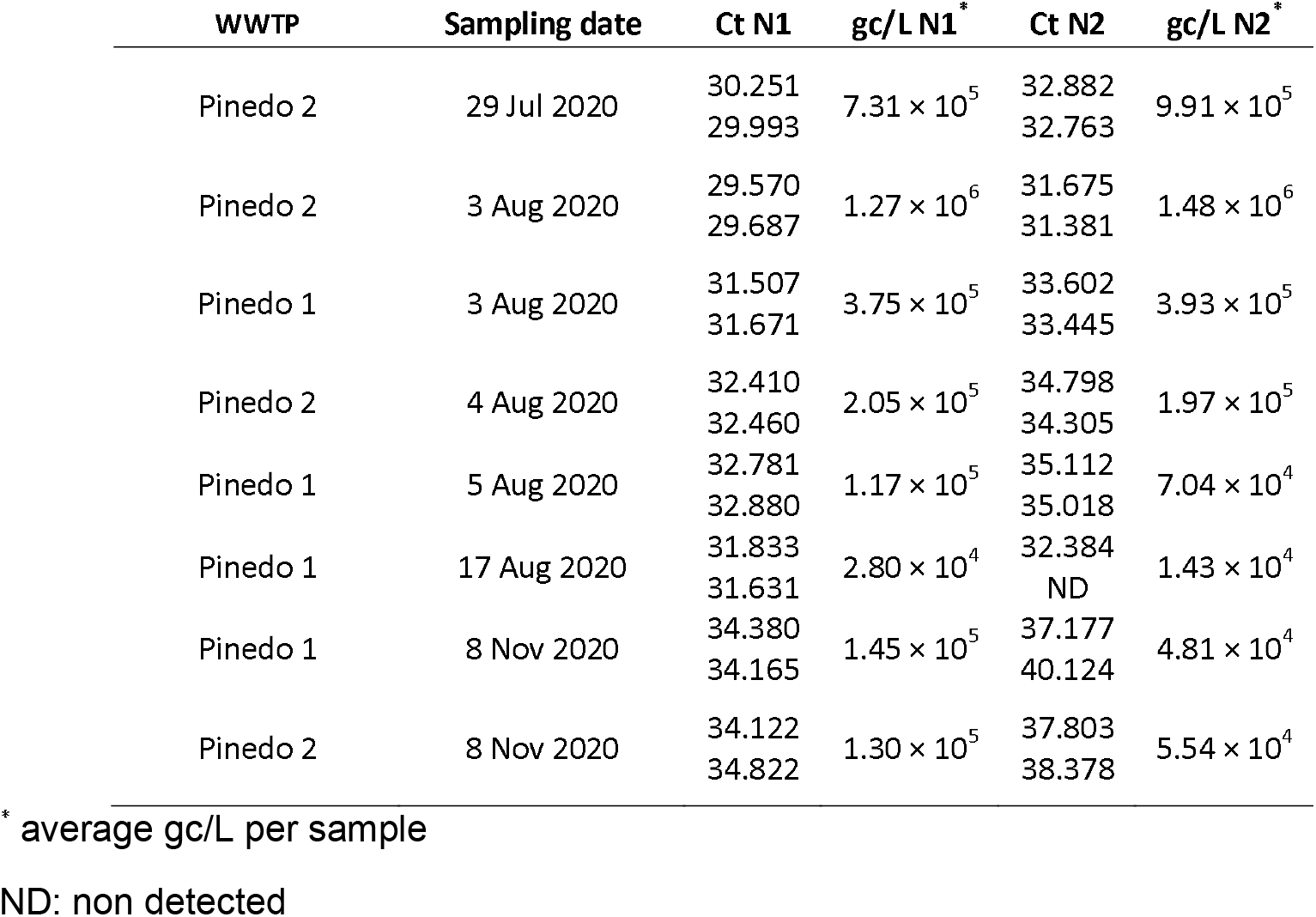
Detection of SARS-CoV-2 RNA in wastewater by RT-qPCR.

**Table 5.**
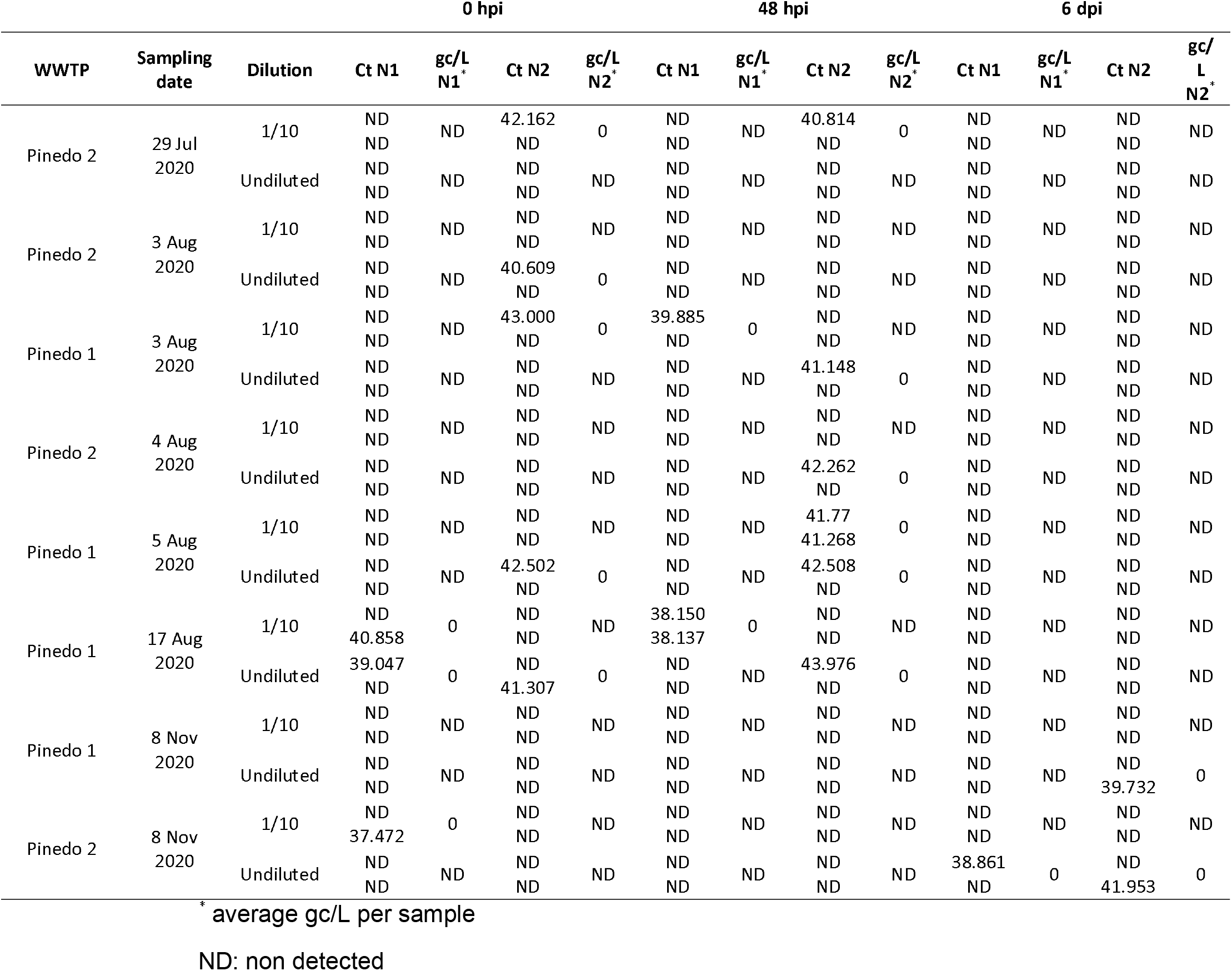
Detection of SARS-CoV-2 RNA in inoculated Vero E6 cultures with concentrated wastewater by RT-qPCR after 0 hpi, 48 hpi and 6 dpi.

## Discussion

Fecal shedding of SARS-CoV-2 has been reported in patients with COVID-19 and in asymptomatic infected individuals. However, the possible fecal-oral transmission route of the coronavirus is still under discussion. Our results are in agreement with those previously published on the detection of SARS-CoV-2 in the stool of COVID-19 patients [20]. However, we were able to detect viral RNA in stool in 6/8 patients, being a high percentage compared to previously published results, and suggesting that the virus may be detected more frequently in stool than expected based on previously published results [21]. Furthermore, our results suggest that GI symptoms and the presence of SARS-CoV-2 in feces are not correlated, as only 3 patients had GI symptoms. This result could have implications for the use of anal swabs as recently suggested. However, the implementation of more sensitive detection methods associated with fecal samples could be an interesting solution to develop non-invasive easy-to-use, rapid tests, than can be performed by common users, avoiding nasopharyngeal swabs, since nasal self-swabbing can lead to inadequate sample collection resulting in false negative conclusions. Thus, the results obtained in this study, despite the low number of patients included, suggest that the presence of SARS-CoV-2 in feces is common, and support the use of anal testing for its detection.

To unravel the fecal-oral route of SARS-CoV-2 transmission, fecal samples have been analyzed in cell culture for the presence of infectious viral particles. The lack of evidence of infectivity in fecal samples could be due to the low viral load found in our samples, with high Ct values, in contrast with previously published results in which infectious coronavirus was found in a sample with a Ct value ranging 20-23 [11]. The fact that only a few cases of infectious particles in feces have been reported so far suggests that although transmission through feces might exists, it should not be considered as a major route. To go further, sewage samples at the time of highest incidence in Valencia (Spain) were tested in cell culture. Despite the high viral load found in wastewater, no viral replication was detected in the samples tested, suggesting that transmission of SARS-CoV-2 through stool or sewage should be minor or absent. In agreement with our results, a recent study in China was unable to detect viable virus in hospital wastewater [22].

We strongly suggest increased effort to continue testing more patients, including asymptomatic patients, to get a broader view of viral shedding in feces. Further evidence should be conducted in a single-center study involving a large cohort to draw supported conclusions. In addition, sewage sampled directly from sewers could be an interesting approach to detect possible infectious particles, avoiding the wastewater treatment plants, where viral particles could be inactivated due to external factors such as temperature, humidity, or chemical agents in the matrix, which may remove or impact the envelope.

Finally, we used RT-qPCR for RNA detection following the CDC recommendations, and the recommended cellular model for SARS-CoV-2 infection. However, improved detection methodologies using more sensitive and efficient techniques, both in molecular diagnostic and cell culture, could lead to different results. Therefore, further characterization of fecal and sewage samples is mandatory to unravel the possible fecal-oral route of SARS-CoV-2 transmission, and will be an interesting topic for future research, especially with the emergence of new viral variants.

## Data Availability

The data that support the findings of this study are available on request from the corresponding author.

## Acknowledgements

We thank to M. Carmen Montaner from the Hospital Universitari i Politècnic La Fe for her support in the collection of the samples. We thank the Generalitat Valenciana and the Empresa Pública de Saneamiento de Aguas Residuales (EPSAR) for providing access to wastewater. We also want thank Dr. Antonio Alcamí for the Vero E6 cells. This research was funded by FONDO-COVID19 COV20/00210 funded by Instituto de Salud Carlos III to P.D-C. P.D-C. was supported by a Ramón y Cajal contract from the Spanish Ministry of Science and Innovation, Call 2019.

## Declarations

### Availability of data and material

The data that support the findings of this study are available from the corresponding author, upon reasonable request.

### Code availability

Not applicable.

### Ethics approval

This study has been approved by the ethics committee of the Instituto de Investigación Sanitaria La Fe (Valencia, Spain).

### Consent to participate

Informed consent was obtained from all individual participants included in the study.

### Consent for publication

The authors declare consent for publication.

